# Antenatal Education for Labour and Postpartum Pain: A Scoping Review of Content, Approaches, and Gaps

**DOI:** 10.1101/2025.08.03.25332898

**Authors:** Elliot Sloyan, Elizabeth Leddy, Carol Clark, Sinéad Dufour, Rosie Harper, Ömer Elma

**Affiliations:** Department of Rehabilitation and Sport Science, Faculty of Health and Social Sciences, Bournemouth University, Bournemouth, United Kingdom; School of Rehabilitation Science, Faculty of Health Sciences, McMaster University, Ontario, Canada; Universtiy Hospitals Dorset NHS Foundation Trust, Poole, Dorset, United Kingdom; Pain in Motion Research Group (PAIN), Department of Physiotherapy, Human Physiology and Anatomy, Faculty of Physical Education and Physiotherapy, Vrije Universiteit Brussel, Brussels, Belgium

**Keywords:** Antenatal Education, Pain Management, Labour, Postpartum, Scoping Review

## Abstract

**Background:** Pain management during labour and the postpartum period is a critical aspect of maternal care, yet many individuals face challenges in obtaining and implementing effective strategies. Antenatal education programmes play a key role in preparing expectant parents for childbirth and beyond, but their impact on women’s pain management remains unclear. Therefore, this review aims to explore and map the contents and characteristics of existing antenatal education programmes that address labour and postpartum pain, which could inform future efforts to develop and optimise these programmes.

**Methods:** This review followed the PRISMA-ScR and Joanna Briggs Institute guidelines. The protocol was registered with the Open Science Framework (6597j). 11 electronic databases were systematically searched in November 2024. Eligible studies were screened independently by two reviewers using Rayyan software. Inclusion criteria focussed on quantitative trials of antenatal education programmes addressing labour and postpartum pain outcomes. Data were extracted and a narrative synthesis was completed to map intervention characteristics and outcomes.

**Results:** A total of 5,876 records were identified from the search strategy. A total of 12 articles met the eligibility and inclusion criteria, including seven randomised controlled trials and five quasi-experimental studies. The content and structure of antenatal education interventions between studies was heterogenous. Common themes included the distinction between “true and false labour pain” and breathing exercises. Face-to-face delivery of antenatal education was the preferred delivery method.

**Conclusion:** Antenatal education programmes contain limited information on labour and postpartum pain management, with little consistency across interventions. Non-pharmacological pain management strategies have the potential to address biopsychosocial factors of pain and support coping in labour. We recommend the development of interventions based on pain science education principles to support pain-management during labour and in the postpartum.

## Introduction

Women’s experience of pain during labour and the postpartum period is a complex and multifactorial experience influenced by physiological, psychosocial, and cultural factors [1]. Although labour pain is often functional and indicative of progress, it is typically intense and can provoke fear, loss of control, and emotional distress [2]. When poorly managed, labour pain is associated with a range of adverse maternal outcomes, including prolonged labour, increased obstetric interventions, postnatal depression, persistent postpartum pain, and birth trauma [3], [4], [5], [6]. As many as 90% of birthing women globally report experiencing pain they describe as “severe” or “very severe” [7]. Pain intensity and suffering are distinct, with suffering reflecting the emotional dimension of pain, while intensity often refers to the magnitude of the sensation [8]. Psychological factors, such as fear of childbirth and pain catastrophising, are linked to heightened pain perception, greater reliance on pharmacological interventions, and a higher incidence of traumatic birth experiences with long term impact [9], [10], [6].

Antenatal education has become a key feature of pregnancy care, providing expectant parents with knowledge and skills to prepare for labour, birth, and early parenthood [11]. These programmes vary in delivery format and content but often include topics such as stages of labour, birth planning, breastfeeding, new-born care, and partner involvement. Whilst pain management strategies are included [12] there are inconsistencies in how pain education is delivered [13], [14]. Recognising the value of preparation for birth, the World Health Organisation (WHO) recommends antenatal education as an essential component of pregnancy care [15]. However, guidance from regulatory bodies such as the National Institute for Health and Care Excellence (NICE) has yet to mandate consistent inclusion of pain education within antenatal care [16]. There remains an absence of standardisation in antenatal education content, with many programmes lacking comprehensive guidance on managing labour and postpartum pain through conservative means such as pain education and mindfulness strategies, despite a growing body of evidence showing the psychological benefits of antenatal education interventions [17], [18].

In the United Kingdom, the absence of national standards or defined curricula results in wide variation in whether, and how, pain is addressed in antenatal education contributing to the “postcode lottery” of care women receive and widening health inequalities [6], [14]. In addition, the variability in educational provision also contributes to unequal birth experiences and potential over-reliance on pharmaceutical interventions, including epidurals and opioids, which carry associated maternal and foetal health risks [5], [19]. This gap limits opportunities to shape maternal expectations, reduce fear, promote self-efficacy, and support informed decision-making around labour pain management.

Overall, understanding the characteristics of current antenatal education could help better understand the role of education on pain management during labour and in postpartum recovery. Enhancing conservative modes of pain management through pregnancy, labour and birth could improve maternal and foetal outcomes. Therefore, the primary aim of this scoping review is to provide a broad overview of the content and characteristics of current antenatal education programmes that address pain management during pregnancy, labour, and the postpartum period. Specifically, the review seeks to identify and describe existing antenatal programmes focused on pain, examine their content, delivery methods, and dosage, and evaluate the pain-related outcomes they target. By doing this, the review will also highlight gaps in the literature to guide future research and inform clinical practice by providing a clearer understanding of how these programmes could utilise the uptake of non-pharmacological pain management strategies.

## Methods

This scoping review was conducted in accordance with the Preferred Reporting Items for Systematic Reviews and Meta-Analyses Extension for Scoping Reviews (PRISMA-ScR) [20], [21]. Resources to help define research questions and aims from the Joanna Briggs Institute (JBI) were utilised [22]. The protocol for this scoping review was registered with the Open Science Framework (6597j) on 11 November 2024 prior to the review being conducted.

### Search strategy

The following electronic bibliographic databases were systematically searched in November 2024: PubMed, MEDLINE, Web of Science, Cochrane CENTRAL, CINAHL, Academic Search, PsycInfo, Education Source, SPORTDiscus, PsycArticles, SocINDEX. Search terms are presented in tabulated form in the Table 1. The full search strategy is presented in Table 2.

**Table 1:** Search terms.

| **PICO Concept** | **Keywords** | **Search Terms** |
| --- | --- | --- |
| Population | Pregnant women |  |
| Intervention | Antenatal education | parent* education OR parent* preparation OR childbirth education OR childbirth preparation OR birth education OR birth preparation OR antenatal education OR prenatal education or antenatal preparation OR prenatal preparation |
| Comparison | With or without |  |
| Outcome measure | Pain | pain |

**Table 2:**
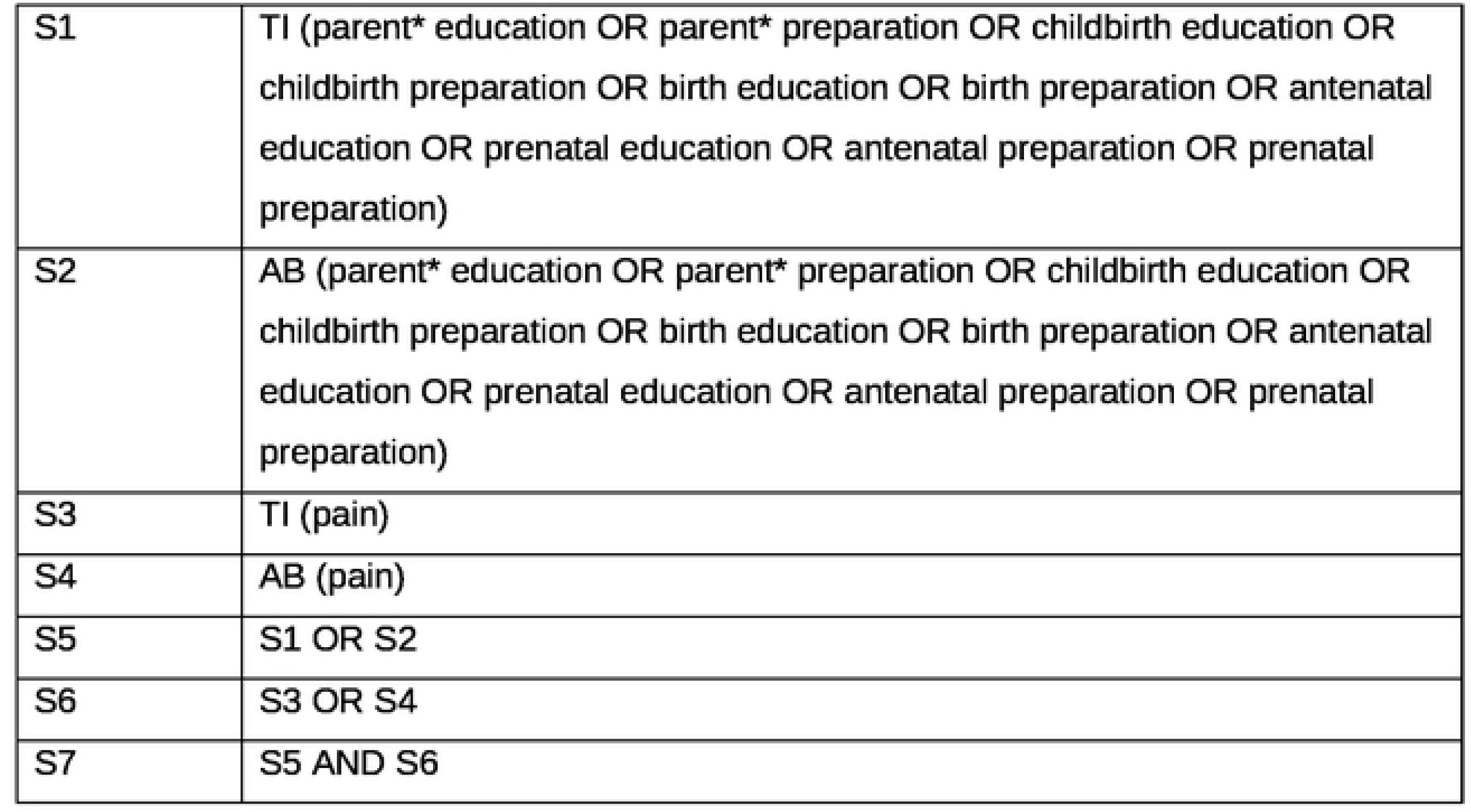
Search strategy.

|  |  |
| --- | --- |
| S1 | TI (parent* education OR parent* preparation OR childbirth education OR childbirth preparation OR birth education OR birth preparation OR antenatal education OR prenatal education OR antenatal preparation OR prenatal preparation) |
| S2 | AB (parent* education OR parent* preparation OR childbirth education OR childbirth preparation OR birth education OR birth preparation OR antenatal education OR prenatal education OR antenatal preparation OR prenatal preparation) |
| S3 | TI (pain) |
| S4 | AB (pain) |
| S5 | S1 OR S2 |
| S6 | S3 OR S4 |
| S7 | S5 AND S6 |

### Study selection and eligibility criteria

The study selection process was completed by two independent and blinded authors (ES and EL) using Rayyan software. Duplicate studies were identified and removed before screening. First, the two authors independently screened all titles and abstracts against eligibility criteria. Then, the full texts of articles that were retrievable were screened independently by the same authors. Discrepancies were resolved through discussion with the third researcher (OE). The search strategy also included forward and backward tracking. Forward tracking involved looking for eligible studies that cited the included studies. Backward tracking involved searching the reference lists of the eligible studies.

No study was excluded based on publication date. This scoping review considered clinical trials of quantitative methodology that explore measurable pain-related outcomes in labour and postpartum among both nulliparous and multiparous women. Trials that explore antenatal education and its effect on perinatal and postnatal pain were included.

The exclusion criteria were as follows: qualitative methodology, protocol studies, pilot studies, feasibility studies, insufficient description of intervention content, male population, outcome unrelated to pain.

### Data extraction and synthesis

Overall, data extraction focused on the contents and characteristics of the antenatal education interventions listed in the captured studies, as well as pain-related outcomes measures. Our data extraction table was developed based on an iteration of a JBI extraction tool, which the screening process helped inform. The extracted data consist of author, year of publication, country, clinical setting, study methodology, key findings, antenatal education content, frequency of education sessions, education delivery method, which profession delivered sessions, pain outcomes measured. Missing or supplementary data was acquired by contacting the study authors as required. The data extraction table is presented in Table 3. Narrative synthesis was employed on included articles based on the aims of the study.

**Table 3:** Data extraction.

| <u>Author;</u><br><u>Year;</u><br><u>Country;</u><br><u>Methodology</u> | <u>Participants</u> | <u>Summary of Intervention and topics</u> | <u>Delivery Method;</u><br><u>Frequency;</u><br><u>Profession</u> | <u>Control Group</u> | <u>Outcome measure</u> | <u>Findings</u> |
| --- | --- | --- | --- | --- | --- | --- |
| Tomyabatra<br>2019<br>Thailand,<br>RCT | Intervention<br>(N = 432)<br>Control<br>(N = 400) | Video of serious complications including pain, vaginal bleeding, water breaking, fewer foetal movements. Distinction between true and false labour pain, maternity role, physical, mental changes, ominous signs to see doctors, sanitation, medication, counting of foetal movement, delivery preparation, alteration of the body near delivery, the distinction between true and false labour pain, abnormal symptoms to see doctors, steps of delivery service in the hospital, a method of delivery, and breathing exercise for labour pain relief | Audio-video media, social networking application, mobile phone<br>Online, mobile phone plus routine ANC<br>"Four times monthly and four times every two weeks"<br>Registered Nurse running ANC | Routine antenatal group-health education<br>Two-time standard ANC group-health education provided by a registered nurse with with information about maternity role, physical, mental changes, ominous signs to see doctors, sanitation, and medication. At 32 weeks' gestation about the counting of foetal movement, delivery preparation, alteration of the body near delivery, the distinction between true and false labour pain, abnormal symptoms to see doctors, steps of delivery service in the hospital, a method of delivery, and breathing exercise for labour pain relief. | **Labour pain duration**<br>Minutes | Significant difference in labour pain duration before admission between the intervention and the control groups (121.7±95.3 versus 139.2±7.0 minutes), coefficient -17.7 (95% CI -31.4 to -4.0); p=0.011. |
| Duncan et al.<br>2017<br>USA<br>RCT | Intervention<br>(N = 15)<br>Control<br>(N = 15) | **The mind in labour (MIL): working with pain in childbirth**<br>Mindfulness strategies for coping with labour-related pain and fear are taught through interactive, experiential activities, with periods of didactic instruction. | F2F<br>18 hours over three days<br>"Mindfulness-Based Childbirth and Parenting Program (MBCP)" instructors | Standard childbirth education courses comparable in length and quality to the MIL intervention but without mindfulness, yoga, or other mind/body components (e.g., hypnosis). A list of approved courses was compiled via a web search and provider follow-ups. If no suitable option was available, participants could submit a preferred course for screening. TAU participants were encouraged to attend with a partner or support person; in this study, 12 reported doing so. | **Retrospect ive perceived labour pain intensity**<br>VAS<br>10cm line<br>"No pain" to "worst possible pain"<br>**Pain catastrophizing scale**<br>Rate degree to which they experience particular thoughts and feelings (e.g., "It's awful and I feel that it overwhelms me" on a scale of 0 (Not at all) to 4 (All the time)) | Condition was marginally associated with pain, with higher scores for MIL participants (average score of 5.20 on the 1-10 VAS) than for control participants (average score of 3.88; b = 1.32, p = .07). When condition was entered in a model with the four covariates, its estimated effect dropped slightly (b = 1.08, p = .14) and the model overall was nonsignificant (Multiple R<sup>2</sup> = .29, p = .15)<br>PCS dropped by 3.6 points in the MIL group and was essentially unchanged in the TAU group (see Fig. 3). The time*group interaction was not significant (t = -1.06, p = .30; estimated treatment effect = -3.26 points, 80% CI [-7.3, 0.8]). When the missing data was imputed, the result did not change (t = -.71, p = .48) |
| Abbasi et al.<br>2017<br>Iran<br>RCT | Intervention (Software) (N = 50)<br>Intervention (Booklet) (N = 51)<br>Control (N = 52) | Both intervention groups had explanation on how to use them, the educational content was explained, and they were asked to study the booklet and software until a sufficient mastery was obtained on the educational content of software and booklet. Participants were called by the researcher every week to remind them to study the training booklet and software.<br>Interventions contained similar content on: situation modification in pregnancy, stretching exercises and breathing, breathing pattern, relaxation, and massage. | F2F<br>One 30-minute session<br>30-34 weeks gestation<br>Midwife | Routine antenatal care and education<br>Not described further. | **Knowledge about pain management**<br>Questionnaire of 10 questions created by researcher. One point for every correct answer. | By adjusting the baseline score, the mean score of knowledge after the intervention in both software group (mean difference = 5.5; CI 95%: 4.6 to 6.3) and booklet group (3.4; 2.5 to 4.2) was significantly higher than the control group. Also, the increase in knowledge score in the software group (2.1; 1.2 to 9.2) was significantly higher than the booklet group |
| Ip et al.<br>2008<br>Hong Kong<br>RCT | Intervention (N = 60)<br>Control (N = 73) | **Self-Efficacy Enhancing Educational Programme (SEEEP) based on Bandura's self-efficacy theory**<br>Demonstration of coping behaviours, including breathing and relaxation, distraction, and cognitive restructuring of pain, to help participants control emotional tensions and pain during labour.<br>Biopsychological phenomena of childbirth; strategies of coping with childbirth discomfort and its relationships with self-efficacy for childbirth. Returning demonstration of the taught coping skills by the participants. Vicarious observation of role models in a VCD depicting how two Chinese pregnant mothers modelled perseverant success in reinstating control over childbirth with the use of coping behaviours. Making a verbal contract to rehearse the taught coping skills with the use of mastery aids at home between and after sessions | F2F group sessions<br>Two 90-minute sessions<br>Nurse | Usual care<br>Physical check-up and attending childbirth classes on voluntary basis. | **Retrospective perceived labour pain intensity**<br>VAS<br>10cm line<br>"no pain at all" to<br>"unimaginable pain" | Intervention group reported reduced perceived labour pain intensity ($p < 0.01$, early stage and $p = 0.01$, middle stage) |
|  |  | Given pamphlet summarising the main coping methods for guiding self-practice and a practice log for daily entries as a means of self-monitoring of their successful practice efforts in the hope that a sense of control was built through structured practice in the exercise of control over challenging conditions<br>Verbal persuasion through positive encouragement. |  |  |  |  |
| Firouzbakht et al.<br>2015<br>Iran<br>Quasi-experimental | Intervention (N = 63)<br>Control (N = 132) | Physical and anatomical changes during pregnancy, psychological health, warning symptoms during pregnancy, the pros and cons of vaginal and caesarean delivery, stages of delivery, breastfeeding, and family planning theoretical training<br>Consultations of 15 min long in forms of questions and answers.<br>Mental and muscular exercises, training proper positions during labour and delivery, proper breathing during pregnancy, labour, and delivery, and 30 min of practicing for pregnant women | F2F plus audio-visual<br>Eight 90-minute sessions<br>Four doulas | Routine prenatal care<br>Not described further. | **Labour pain intensity: collected during labour**<br>VAS 0-100 | Intervention group reported reduced perceived labour pain intensity.<br>Intervention group: 85.68 (1.85)<br>Control group: 90.99 (14.72)<br>(P = 0.03) |
| Mahnaz et al.<br>2019<br>Iran<br>RCT | Intervention (N = 36)<br>Control (N = 34) | Presentation covering all objectives and including photos and posters followed by Q&A.<br>Awareness of physiologic changes in the body during pregnancy, such as breast enlargement, nausea, fatigue, pelvic inflammation, and abdominal and perineal changes, changes in orgasm patterns during sexual activity in this period, cultural and religious restrictions affecting women's sexual function in pregnancy, awareness of sexual function changes in the third trimester of pregnancy, using sexual positions with fewer problems, avoiding sexual activity in case of uterine contractions and vaginal bleeding (i.e., see a doctor as soon as possible, avoiding sexual activity in case of an incompetent cervix, prevention of sexually transmitted diseases<br>In addition, an educational booklet written in simple language was provided for the pregnant women to study at home with their husbands. During the 4 weeks after the last session, the intervention group received no further contact or additional information (other than the cases provided in the educational pamphlet) | F2F group sessions<br>Four sessions<br>Healthcare workers | Training on benefits of breastfeeding and normal delivery.<br>No further description. | **Sexual pain intensity**<br>Included within questionnaire | No difference in sexual pain intensity after intervention (P = 0.78) |
| Ucar and Golbasi<br>2019 | Intervention (N = 52)<br>Control | Diagnosis and evaluation of the perception of childbirth, getting acquainted between educator and participants, configuration of group sessions (Aim, method, duration, | F2F group classes | No intervention / education<br>No further description. | **Retrospective perceived** | Intervention group reported reduced perceived labour pain intensity ((r = -0.421, p = .000) |
| Turkey<br>Quasi-experimental | (N = 59) | frequency, etc.), sharing the beliefs and perceptions of the group members about childbirth, summary<br>Diagnosis of fear of childbirth, feedback on the previous session, setting and sharing the agenda for the session A-B-C model of cognitive behavioural approach, A-B-C model of fear of childbirth, summary<br>Giving "Fear diary" homework, "Severity of labour contractions diary" homework<br>Information about childbirth and correction of misinformation; mood; feedback on previous session; factors that affect childbirth; signs of labour starting; stages of labour; delivery room and the procedures carried out in delivery room; summary<br>Giving "Unfinished stories" homework<br>Coping with uterine contractions; feedback about the previous session; breathing techniques used at the first stage of labour; pushing techniques used at the second stage of labour; summary<br>Giving homework "Unfinished stories" homework; "Birth plan" homework<br>Coping with fear of childbirth and creating a more positive perception of birth; mood; feedback on previous session; reviewing homework; creating a positive perception of birth; relaxation techniques; summary<br>Feedback on previous session; strengthening the breathing techniques used during labour; reviewing information obtained during the group process; evaluation of the group process | Six 45-minute sessions over three weeks<br>Midwives |  | **labour pain intensity**<br>NPRS (VAS)<br>0-10<br>"no pain" to "unbearable pain" |  |
| Camlibel and Mete<br>2023<br>Turkey<br>Quasi-experimental | Intervention (N = 30)<br>Control (N = 30) | Introduction, thoughts, and expectations about the training program, emotions, and thoughts about the concept "delivery", the underlying reasons of positive/negative emotions and thoughts about delivery, history of fear, explaining the aims of the class, "Fear-Tension-Pain" cycle, the role of the hormones in delivery, the effect of fear on the hormones needed for the delivery and on the delivery action, explaining the aims of the preparation for delivery classes, summary, muscle and relaxation exercises, assignment.<br>Summary of previous class, sharing the assignments of the first class, the three rules that are influential in changing the viewpoint on delivery (thought, emotion and behaviour, the power of the language, motivation), the<br>methods that may be used to ensure relaxation (breath exercises, visualisation/imaging, imagination, forming a mental area), muscle and relaxation exercises, assignment.<br>Sharing the assignments of the second class, summary of previous class, the indications of the start of the delivery, real/fake birth pains, the stages of the delivery and its mechanism (opening, delivery, the birth of the placenta), the practices recommended to be made at home when the delivery starts, the hospital process, the delivery video, talking to the doctor and other healthcare staff about the delivery, muscle and relaxation exercises, assignment.<br>Sharing the assignments of the third class, summary of previous class, last preparations for the delivery (delivery pack, transportation to hospital), caesarean, epidural anaesthesia, postpartum early period, delivery video or positive delivery history, relaxation exercises, ceremony for participation certificate, evaluation of the training. | F2F group sessions<br>Four 120-minute sessions total (once weekly for four weeks)<br>Nurses | Routine antenatal care<br>No further description | **Labour pain intensity**<br>Latent phase<br>Active phase<br>Transition phase<br>VAS<br>10cm ruler | Significant difference between the study and the control group in terms of latent phase (U=2.260, p= 0.024), and active phase (U=2.630, p=0.009) pain points averages; while there was no significant difference between the transition phase (U=1.611, p=0.107) pain points averages. |
| Citak Bilgin et al.<br>2019<br>Turkey<br>Quasi-experimental | Intervention (N = 64)<br>Control (N = 57) | Introduction, the aim of the course, anatomy and physiology of reproductive organs, the role of uterus and baby during delivery, physiology of pain in birth, fear and its effects at birth, relaxation and mental imagination techniques, initial symptoms of birth<br>Stages of birth, true and false labour pain, the effect of hormones at birth, emotional and physical support at birth, breathing techniques, communication with the baby in the womb and its importance, effects of relaxation on mother and foetus, progressive relaxation<br>Non-pharmacological techniques used in coping with labour pain, massage techniques, positions can be used at birth, the use of a birthing ball, pushing techniques<br>Anaesthesia at labour, caesarean section, interventions to birth, the establishment of a delivery plan<br>Mother-baby relationship, baby's care (bathing, dressing, etc.), breast milk and its importance, breastfeeding, breastfeeding techniques, breast problems and care, milking with hand and pump, storage of the milk, summary what has been learned in the education and handing out of certificates | F2F<br>Five 3-hour sessions total. (One 3-hour session weekly for five weeks)<br>Doctor of nursing | No intervention / education | **Retrospective perceived labour pain intensity**<br>VAS 0-10 "no pain at all" to "unimaginable pain" | Intervention group reported significantly reduced perceived labour pain intensity ($p = 0.016$) |
|  |  | The participants were given music CDs and education booklets at the beginning of each session and were asked to repeat the exercises with music at home every day. |  |  |  |  |
| Miquelutti et al.<br>2013<br>Brazil<br>RCT | Intervention<br>(N = 97)<br>Control<br>(N = 100) | **Birth Preparation Programme (BPP): physical + educational activities. In addition to routine antenatal care**<br>Guidance on the awareness of pelvic floor muscles—they were instructed to contract the pelvic floor as if they were trying to avoid urination and focus on the sensation of the pelvic floor muscles during this movement and during relaxation after the contraction. Women who did not manage to perform this exercise were advised to try the contraction of the pelvic floor during urination, attempting to stop urine flow and informed that this exercise during urination was only for the identification of the sensation. During BPP meetings information was provided on the prevention of pain in pregnancy, the role of the pelvic floor in pregnancy, delivery and in the postpartum, the physiology of labour, breathing exercises for delivery, and non-pharmacological pain control techniques during labour. Participants received a guide with the exercises to be performed daily at home, consisting of: pelvic floor muscle training (PFMT) including rapid (30 times) and sustained maximal contractions (20 times holding for 10 seconds), stretching exercises to reduce back pain and exercises to improve venous return in the lower limbs. The women were also encouraged to practice aerobic exercise daily for at least 30 minutes and received written information based on the ACOG guidelines regarding the warning signs that indicated that the exercise should be stopped<br>Short description of the BPP exercises<br>• Stretching: Head and neck; anterior, posterior, and lateral trunk; lower limbs; active mobilisation of the spine and pelvis in the position of all fours; lumbar traction<br>• Venous return: Exercises with the lower limbs in the standing and lateral decubitus position<br>• Abdominal exercises: Transverse abdominal muscle activation in both standing and all four positions<br>• PFMT: Maximal rapid and sustained contraction on standing and sitting position<br>• Relaxation: Training of breathing techniques for contraction control during labour; progressive relaxation techniques; massage; mentalisation | F2F<br>50-minutes, fortnightly between 31 and 36 weeks of pregnancy and weekly from 37 weeks of pregnancy onwards<br>Physiotherapists, Nurses, Medical Staff | Routine antenatal care<br>Educational activities routinely offered at the prenatal clinics: information on breastfeeding, the signs and symptoms of labour and a visit to the delivery ward. During labour, at the maternity ward, non-systematic information on the use of non-pharmacological pain relief techniques were provided by trained physiotherapy, nursing, and medical staff. | **Lumbopelvic pain**<br>VAS<br>10cm ruler | No difference in lumbopelvic found between groups |
| Heim and Makuch<br>2024<br>Brazil<br>Quasi-experimental | Intervention (N = 50)<br>Control (N = 50) | All educational sessions were similar when performed in-person or virtually (by WhatsApp platform) and began with a brief conversation to encourage women to share their feelings, doubts and concerns about labour and delivery.<br>Topics: contractions, pain during labour and delivery, and how to seek relief by adopting the upright position and using breathing techniques and relaxation. The non-pharmacological techniques were selected because they constitute a complementary approach to pain relief. Once women learn and become confident with their use, they become independent and more active in controlling their birthing process, they can be used by the woman regardless of other resources and are harmless for the baby and the mother. Breathing techniques, upright positions and relaxation techniques were performed by the pregnant women at all educational meetings. For the breathing technique, women were instructed to take a deep, nasal, slow abdominal inspiration, followed by a deep, prolonged exhalation through the mouth. The women were stimulated to maintain simultaneously global relaxation in the chosen position, mainly an upright position ('smell the flower and blow out the candle flame at the same time relaxing your body, feel where you are tense, and relax your body'). Among the vertical and comfortable positions during labour, the following positions were trained: sitting on a birthing ball, standing, walking between contractions, squatting, and sitting, and any vertical position that women felt was comfortable and helped to control pain, to perform the breathing techniques and to relax. All the exercises were trained during a time like the duration of a contraction. | F2F + virtually (via WhatsApp)<br>Two-five 20-minute sessions<br>Interrupted by COVID-19 pandemic<br>Physiotherapists, Nurses, Physicians | No intervention / education | **Retrospective perceived labour pain intensity**<br>VAS<br>0-10<br>"no pain" to "maximum level of pain tolerated" | No difference in intensity labour pain reported between the two groups<br>Intervention group: 8.8 (SD=0.23)<br>Control group: and 9.1 (SD=0.29) (p<0.03) |
| **Backwards-forwards**<br>El-Kurdy et al.<br>2017<br>Egypt<br>RCT | Intervention (N = 52)<br>Control (N = 52) | Overview of Labour and Delivery:<br>Welcome & develop rapport among group participants; explain objectives; encourage participants to express their feeling and sharing thoughts about childbirth process; introduction to pre-test questionnaires (Interview – CBSEI); anatomy and physiology of female genital organs; premonitory signs of labour; true and false labour pain; what to bring to the hospital; stages and phases of labour<br>Coping with Labour Pain:<br>Review content from class one; nature of labour pain; non-pharmacological coping measures with labour pain; medications used in labour<br>Delivery and Postpartum: | F2F<br>Posters, slide presentations, animation videos, and demonstration<br>Three 90-minute weekly sessions<br>Midwives | Routine ANC<br>Exposed to all conditions of intervention group except for antenatal education sessions. | **Retrospective perceived labour pain intensity**<br>VAS<br>0-10<br>"no pain" to "most intense pain" | 1st stage: antenatal education and control groups respectively (5.08 ± 0.68 & 7.40 ± 0.5).<br>2nd stage: antenatal education and control groups respectively (6.52 ± 0.5 & 8.56 ± 0.7).<br>Highly significant differences between the two groups regarding the mean score of labour pain at the 1st and 2nd stages of labour (p = <0.001). |
|  |  | Brief review the content of class 1-2 and ask for the question and discuss the answer if they have; delivery variations (Episiotomy and C-section); immediate postpartum care in hospital; new-born care; post-test (CBSEI) |  |  |  |  |

ANC = antenatal care
F2F = face-to-face
NPRS = numerical pain rating scale
RCT = randomised controlled trial
VAS = visual analogue scale

### Quality appraisal

The methodological quality of the included studies was assessed using the Downs and Black checklist [23], as represented in Table 4. This checklist comprises 27 questions and examines the reporting, external validity, internal validity, and power of an interventional study [23]. While scoping reviews do not typically require quality appraisal, its inclusion in this review helps improve synthesis and increase the practical utility of the research findings. Further, the inclusion of quality appraisal enhances transparency and provide essential context for interpreting the evidence. This is especially relevant as the findings may inform future experimental studies and clinical applications.

**Table 4:** Methodological quality appraisal (Downs and Black)

|  |  | Tomyabatr<br>a 2019 | Duncan<br>et al.<br>2017 | Abbasi<br>et al.<br>2017 | Ip et al.<br>2008 | Firouzbakht<br>et al. 2015 | Mahnaz<br>et al.<br>2019 | Ucar and<br>Golbasi<br>2019 | Camlibel<br>and Mete<br>2023 | Citak<br>Bilgin et<br>al. 2019 | Miquelutti<br>et al. 2013 | Heim and<br>Makuch<br>2024 | El-Kurdy<br>et al. 2017 |
| --- | --- | --- | --- | --- | --- | --- | --- | --- | --- | --- | --- | --- | --- |
| **Reporting** |  |  |  |  |  |  |  |  |  |  |  |  |  |
| 1 | Is the hypothesis/aim/objective of the study clearly described? | 1 | 1 | 1 | 1 | 1 | 1 | 1 | 1 | 1 | 1 | 1 | 1 |
| 2 | Are the main outcomes to be measured clearly described in the Introduction or Methods section? | 1 | 1 | 1 | 1 | 1 | 1 | 1 | 1 | 1 | 1 | 1 | 1 |
| 3 | Are the characteristics of the patients included in the study clearly described? | 1 | 1 | 1 | 1 | 1 | 1 | 1 | 1 | 1 | 1 | 1 | 1 |
| 4 | Are the interventions of interest clearly described? | 1 | 1 | 1 | 1 | 1 | 1 | 1 | 1 | 1 | 1 | 1 | 1 |
| 5 | Are the distributions of principal confounders in each group of subjects to be compared clearly described? | Partial | 0 | 0 | Partial | 0 | 0 | Partial | 0 | 0 | 1 | 0 | 0 |
| 6 | Are the main findings of the study clearly described? | 1 | 1 | 1 | 1 | 1 | 1 | 1 | 1 | 1 | 1 | 1 | 1 |
| 7 | Does the study provide estimates of the random variability in the data for the main outcomes? | 1 | 1 | 1 | 1 | 1 | 1 | 1 | 1 | 1 | 1 | 1 | 1 |
| 8 | Have all important adverse events that may be a consequence of the intervention been reported? | Partial | 0 | 0 | 0 | 0 | 0 | 0 | 0 | 0 | 0 | 0 | 0 |
| 9 | Have the characteristics of patients lost to follow-up been described? | Partial | Partial | 1 | 1 | 0 | 1 | 1 | 1 | 1 | 1 | 1 | 1 |
| 10 | Have actual probability values been reported? | 1 | 1 | 1 | 1 | 1 | 1 | 1 | 1 | 1 | 1 | 1 | 1 |
| **External Validity** |  |  |  |  |  |  |  |  |  |  |  |  |  |
| 11 | Were the subjects asked to participate in the study representative of the entire population from which they were recruited? | UTD | UTD | UTD | UTD | UTD | UTD | UTD | UTD | UTD | UTD | UTD | UTD |
| 12 | Were those subjects who were prepared to participate representative of the entire population from which they were recruited? | UTD | UTD | UTD | UTD | UTD | UTD | UTD | UTD | UTD | UTD | UTD | UTD |
| 13 | Were the staff, places, and facilities where the patients were treated, representative of the treatment the majority of patients receive? | 0 | 0 | 0 | 0 | 0 | 0 | 0 | 0 | 0 | 0 | 0 | 0 |
| **Internal Validity – Bias & Confounding** |  |  |  |  |  |  |  |  |  |  |  |  |  |
| 14 | Was an attempt made to blind study subjects to the intervention they have received? | 0 | 0 | 0 | 0 | 0 | 0 | 0 | 0 | 0 | 0 | 0 | 0 |
| 15 | Was an attempt made to blind those measuring the main outcomes of the intervention? | 0 | UTD | 0 | 1 | 0 | 1 | 0 | 0 | 0 | 0 | 0 | 0 |
| 16 | If any of the results of the study were based on "data dredging", was this made clear? | UTD | 1 | 0 | 1 | 0 | 1 | 1 | 1 | 1 | 1 | 1 | 1 |
| 17 | In trials and cohort studies, do the analyses adjust for different lengths of follow-up of patients, or in case-control studies, is the time period between the intervention and outcome the same for cases and controls? | 1 | 1 | 1 | 1 | 1 | 1 | 1 | 1 | 1 | 1 | 1 | 1 |
| 18 | Were the statistical tests used to assess the main outcomes appropriate? | 1 | 1 | 1 | 1 | 1 | 1 | 1 | 1 | 1 | 1 | 1 | 1 |
| 19 | Was compliance with the intervention/s reliable? | Partial | 1 | Partial | Partial | 0 | Partial | Partial | 0 | 0 | 0 | 0 | Partial |
| 20 | Were the main outcome measures used accurate (valid and reliable)? | 1 | 1 | Partial | 1 | 1 | 1 | 1 | 1 | 1 | 1 | 1 | 1 |
| 21 | Were the patients in different intervention groups (trials and cohort studies) or were the cases and controls (case-control studies) recruited from the same population? | 1 | 1 | 1 | 1 | 1 | 1 | 1 | 1 | 1 | 1 | 1 | 1 |
| 22 | Were study subjects in different intervention groups (trials and cohort studies) or were the cases and controls (case-control studies) recruited over the same period of time? | 1 | 1 | 1 | 1 | 1 | 1 | 0 | 0 | 0 | 1 | 0 | 1 |
| 23 | Were study subjects randomised to intervention groups? | 1 | 1 | 1 | 1 | 0 | 1 | 0 | 0 | 0 | 1 | 0 | 1 |
| 24 | Was the randomised intervention assignment concealed from both patients and health care staff until recruitment was complete and irrevocable? | 1 | 1 | 1 | 0 | 0 | Partial | 0 | 0 | 0 | 1 | 0 | 0 |
| 25 | Was there adequate adjustment for confounding in the analyses from which the main findings were drawn? | 0 | 0 | Partial | Partial | Partial | 0 | 0 | 0 | 0 | 0 | Partial | 0 |
| 26 | Were losses of patients to follow-up taken into account? | 0 | Partial | 1 | 0 | Partial | 1 | Partial | Partial | 0 | Partial | 0 | 0 |
| **Power** |  |  |  |  |  |  |  |  |  |  |  |  |  |
| 27 | Did the study have sufficient power to detect a clinically important effect where the probability value for a difference being due to chance is less than 5%? | 0 | 0 | 0 | 1 | 0 | 1 | 1 | 1 | 1 | 1 | 1 | 1 |
| **Total Score** |  | **14** | **16** | **15** | **17** | **13** | **18** | **14** | **14** | **14** | **17** | **14** | **16** |

UTD = unable to determine

## Results

### Study selection and characteristics of included studies

The details of the process of selecting studies in accordance with PRISMA-ScR standards can be found in Figure 1. A total of 5,876 studies were retrieved and screened against the inclusion and exclusion criteria. A total of 12 studies met the inclusion criteria, including seven Randomised Controlled Trials (RCTs) [24], [25], [26], [27], [28], [29], [30] and five quasi-experimental studies [31], [32], [33], [34], [35].

**Figure 1:**
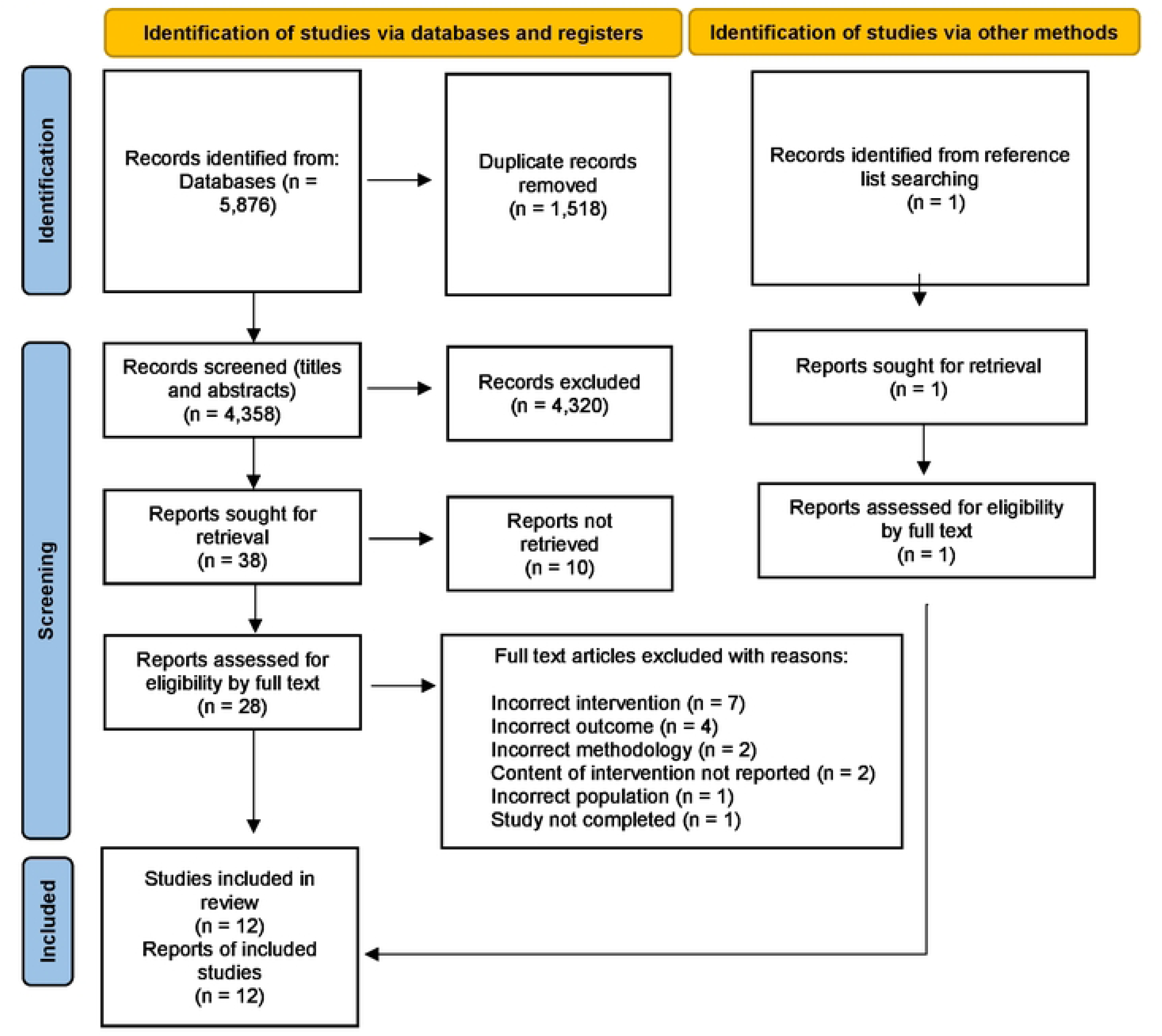
PRISMA flow diagram

The included studies were published between 2008 and 2024 and spanned populations from seven countries: Iran, Turkey, Thailand, Egypt, Hong Kong, Brazil, and the United States. Sample sizes ranged from 30 [27] to 832 [30], with a total of 2,106 participants. A comprehensive overview of the characteristics of the selected studies can be found in Table 3.

### Mode of antenatal education

Among the included studies, antenatal education was delivered in three of the following way: face to face, a hybrid of in person and online, and online only. Nine studies were solely face-to-face [24], [25], [26], [27], [28], [32], [29], [33], [34]. Two studies used a combination of face-to-face and virtual online materials [31], [35]. One study utilised virtual online materials only which included audio-video media through a social networking application [30].

### Frequency and duration of antenatal education

The frequency and duration of antenatal education sessions varied widely across all included studies. The length of the sessions varied from 20 minutes [35] to six hours [27]. Total time spent in antenatal education sessions ranged from 30 minutes [26], to 18 hours [27]. Two studies reported the same total time spent on antenatal education, 4.5 hours [28], [33], while all remaining studies differed in total time [24], [25], [31], [26], [27], [32], [29], [30], [34], [35]. Three studies reported their sessions were 90 minutes in length [24], [31], [28]. The number of antenatal education sessions ranged from one [26], to eight sessions [31], [30]. Frequency of sessions ranged from daily for three days [27] to fortnightly [25].

### Profession(s) delivering the antenatal education

In 10 studies, only one profession delivered the antenatal education [24], [31], [26], [27], [28], [32], [29], [30], [33], [34], whereas in the other two studies, a combination of professions delivered the education [25], [35].

Nurses delivered antenatal education in three studies [24], [30], [34], as did midwives [26], [28], [33]. Doulas [31], a Doctor of Nursing [32], ‘mindfulness instructors’ [27] and ‘healthcare workers’ [29], each delivered the intervention in one study respectively. A combination of physiotherapists, nurses, and physicians delivered the education in one study [35], while a combination of physiotherapists, nurses, and ‘medical staff’ delivered education in one study [25].

### Control group among included studies

All 12 studies included in the review had a control group. The control groups in eight of the reviewed studies received routine antenatal care [24], [25], [31], [26], [27], [28], [30], [34]. Three control groups received no education nor intervention [32], [33], [35]. Mostly, control participants attended general antenatal sessions that covered basic pregnancy care, foetal development, and signs of labour, but did not specifically focus on pain management techniques [30], [28]. Participants in the control group of one study received training on the benefits of breastfeeding and normal delivery [29].

### Pain outcome measures

Eight of the 12 included studies measured labour pain intensity using a visual analogue scale (VAS) [24], [31], [27], [28], [32], [33], [34], [35]. Seven of these studies measured labour pain retrospectively [24], [31], [27], [28], [32], [33], [35], while one measured during labour [34]. Additional outcome measures used included: labour pain duration [30]; knowledge on pain management using a self-produced questionnaire [26]; sexual pain intensity postpartum [29]; pain catastrophising scale in labour [27]. One study measured postpartum pain, namely dyspareunia [29].

### Content of antenatal education packages targeting pain

Of the 12 included studies, interventions were described to varying levels of detail. Generic and varied pain-related topics were listed in eight studies [24], [25], [27], [28], [32], [30], [34], [35], whilst four did not mention pain [31], [26], [29], [33]. Only one of 12 studies explicitly stated a proposed mechanism by which their intervention addressed pain [30],, proposing their birth preparation program “may reduce anxiety and fear of childbirth and increase self-efficacy and confidence by offering knowledge and practical coping skills. This may influence the perception of labour pain and enhance birth experience and satisfaction” [30].

Antenatal education interventions covered a range of pain-related topics aimed at preparing pregnant women for labour and postpartum pain management. Four interventions included the distinction between “true and false labour pain”, helping participants recognise the signs of active labour [28], [30], [32], [34]. Only one study reported including education on pain physiology [32]. Cognitive restructuring and self-efficacy training were also introduced in some programmes [24], [33].

Non-pharmacological pain management techniques were commonly included in conjunction with antenatal education, such as breathing techniques, relaxation techniques, pelvic floor exercises, massage, and a birthing ball. Six studies referred to breathing exercises and relaxation techniques within their intervention to help manage labour pain [30], [26], [31], [33], [34], [32], [25], [35]. Three studies reported utilising massage [25], [26], [32]. Two studies reported using a birthing ball [32], [35]. One study reported teaching pelvic floor exercises to facilitate pain reduction and improve comfort during labour [25]. No study mentioned the use of transcutaneous electrical nerve stimulation (TENS).

### Quality appraisal

The methodology quality scores based on the Downs and Black checklist are presented in Table 4. Scores ranged between 13 [31] and 18 [29]. The mean score was 15. According to the checklist, an average score of 15 means the quality of the included studies was fair, with some methodological limitations. No study was able to blind participants to the intervention they were receiving. One study partially explained adverse events [30], while the remaining studies did not. When assessing internal validity, two studies reported blinding those measuring the main outcomes [24], [29], while no study was able to blind study subjects to the intervention they had received. External validity was universally limited as the researchers were unable to determine if study participants were representative of the entire population. Similarly, they were unable to determine if the staff and facilities were representative of the treatment the majority of patients receive.

## Discussion

This scoping review aimed to map the contents and characteristics and gaps of current antenatal education programmes that target pain management during pregnancy, labour, and the postpartum period. In doing so, it aimed to highlight gaps in the literature to inform future research and enhance clinical practice, ultimately improving maternal and neonatal health outcomes while reducing strain on healthcare systems. This scoping review identified 12 studies exploring antenatal education programmes targeting labour and postpartum pain, published between 2008 and 2024 across seven countries. Most commonly, methodological quality of the included studies was considered fair. Distinguishing between true and false labour pain was the most reported topic within the included interventions. Most studies employed face-to-face delivery, with a minority using virtual or hybrid formats. There was substantial variability in session frequency, duration, and total exposure time. Education was most often delivered by a single professional, though multidisciplinary approaches were also noted. Control groups primarily received routine antenatal care, with limited focus on pain-specific education. Eight studies measured labour pain intensity using a VAS, predominantly retrospectively. Only one study articulated a clear mechanism linking education to pain outcomes, and only one study incorporated elements of pain neuroscience education. Techniques such as breathing, relaxation, massage, and cognitive strategies were used inconsistently across interventions.

### Themes

Despite each of the included studies measuring a pain-specific outcome, only eight of the 12 studies mentioned pain when describing the intervention. Furthermore, none of these eight studies provided a detailed description of the education content specific to pain-management such that the programme could be replicated. Descriptions of the interventions included: “information was provided on the prevention of pain in pregnancy” [25]; “nature of labour pain” [28]; “Fear-Tension-Pain cycle” [34]. This lack of detail suggests either a limited depth in understanding of pain-related topics, absent consensus about which pain-related topics should be prioritised within antenatal care, or both. Antenatal education services remain largely unregulated, and academics and clinicians in this area appear to share the view that these services should be standardised across healthcare settings [36], [14]. Findings from this review reinforce the need for consistent, pain-specific content within antenatal education programmes. This may support the development of reproducible interventions and contribute to the optimisation and standardisation of pain-focused antenatal education across antenatal settings to improve maternal outcomes.

The topic of distinguishing between “true and false labour pain” was reported in four of the included studies. True labour pain may be defined as regular, increasing uterine contractions that cause progressive cervical dilation and lead to childbirth, whereas false labour pain can be categorised as irregular, non-progressive contractions that do not cause cervical changes and do not result in labour. This is important as false recognition of labour can have many unwanted consequences. Firstly, mistaking false labour for true labour can increase fear, particularly for nulliparous women, which may contribute to overusing obstetric interventions [37]. Heightened fear of childbirth is identified as a common reason for caesarean delivery [38], and thus a deterrent from pre-existing plans for physiological birthing. Furthermore, unnecessary hospital visits may place additional strain on midwives and the health system, potentially detracting from their care of women in true labour [39]. These reasons may partially explain the emphasis of this topic within the included interventions. Conversely, there was limited focus of antenatal education on pain self-management techniques during labour. NICE guidelines [40] recommend the use of water, TENS, and music during intrapartum care, however none of these interventions were discussed in the included studies. Future experimental studies exploring the effect of antenatal education should include the topic of distinguishing between true and false labour pain and signs alongside conservative pain-management options.

This review found an absence of explicitly stated mechanisms by which pain was managed. Similarly, only one study reported including the topic of pain physiology. The inferred mechanisms of the remaining studies were mostly psychological, for example, increasing self-efficacy can improve coping, and reducing fear and anxiety can lower pain perception. Contemporary understanding of pain science has significantly developed in the last two decades. The recency of such developments, and a growing dependence on medicalised birth [41], may suggest this knowledge of pain has not yet been understood, adopted, nor incorporated by midwives and other professionals involved in labour pain-management. This may help explain the absence of pain physiology in the included studies, as well as the lack of proposed mechanisms of action. Understanding the neurophysiological mechanisms of pain modulation has proven effective in reducing rates of medicalised birth: in their meta-analysis of nonpharmacologic approaches for labour pain management, Chaillet et al. [11] highlighted an association between interventions based on central nervous system control, including pain education, significantly reduced rates of caesarean delivery and pain medication usage. This prompted the 2018 Canadian Obstetric Guideline No. 355 that recommended educating staff on neurophysiology of pain perception to increase physiological birth rates [42]. These findings highlight the need for a greater emphasis on understanding the neurophysiological mechanisms of the pain experience during labour and the postpartum period.

Of the included studies, seven explored labour pain intensity with a version of the VAS, while only one included a postpartum pain-outcome, dyspareunia. The VAS is a psychometric measure of subjective pain experience used widely within research and clinical practice [43]. Among other benefits, the VAS is supported for its simplicity and versatility. However, the VAS reduces pain to a unidimensional experience, which conflicts with modern understanding of pain as a complex and multidimensional phenomenon [44]. Further, most studies captured pain retrospectively that influenced the reported scores. Future studies may look to incorporate outcome measures that better capture the qualitative components of the labour pain experience [45].

Duncan et al. [27] were the only researchers to use the pain catastrophising scale as an outcome measure. This outcome appears underrepresented based on its proven importance in predicting long-term postpartum health. Pain catastrophising behaviour throughout pregnancy has consistently been associated with poor labour and postpartum outcomes. These include perineal trauma and persistent perineal pain following vaginal delivery [46], [47]; postpartum depression; poor social adjustment after delivery [48]. These emotional and social aspects of postpartum health are important as they have been shown to contribute significantly to a woman’s experience of pain [49] alongside socioeconomic status and culture backgrounds and beliefs [40]. Increasing women’s knowledge of pain has been shown to reduce pain catastrophising scores [50]. Interventions utilising pain neuroscience education during pregnancy have shown improvements in pain knowledge, birth self-efficacy, labour coping and lower rates of caesarean delivery and epidural use [51]. [52]. Therefore, to target pain catastrophising and other positive birth outcomes, it is recommended that future experimental studies develop and refine pain neuroscience education for labour and postpartum pain management.

The overarching delivery method of antenatal education across the included studies was face-to-face. In-person clinics often include physical examinations alongside education, which cannot be completed virtually. Primiparous women across cultural and socio-economic backgrounds report a preference for in-person sessions to facilitate asking questions and connecting with other expectant parents [53]. However, the COVID-19 pandemic initiated a cultural shift towards telehealth, with its use increasingly prevalent [54]. An integrative review of women’s experiences of online antenatal education identified flexibility as a key benefit, accommodating geographical barriers, illness, and complex social situations [55]. Additional advantages included enhanced comprehensibility, the ability to revisit content, timely information delivery, and broader accessibility. Despite this, those with lower e-health literacy may face difficulties accessing digital information, contributing to a ‘digital divide’ [55]. Pregnant women are known information seekers, yet challenges exist regarding the trustworthiness of online sources [56], [57], with many women citing difficulty distinguishing reliable from false pregnancy and labour information [58]. When reliable, evidence-based information is used and delivered effectively, digital education has been shown to play an important role in antenatal care. A recent study evaluated a mobile application that included pain neuroscience education combined with mindfulness training to prepare for birth [52]. A large sample size was studied and improved pain knowledge, improved self-efficacy in birth as well as lower rates of caesarean birth and epidural use were demonstrated as a result of the intervention. Such findings considered alongside two studies included in this review which used a hybrid approach, that included online resources, point to the potential innovative digital modes of care provision provided quality control measures to ensure rigor are fulfilled.

There was wide disparity in the frequency and duration of antenatal education interventions, ranging from one 30-minute session to five three-hour sessions. Pain-specific content formed only a portion of each session, meaning actual time spent on pain education was less than stated. No included country currently has mandated recommendations on the number, frequency, duration, or timing of antenatal sessions. This heterogeneity may reflect that clinical delivery is not meeting current guidelines based on service pressures [59]. While any antenatal education was linked to reduced pain medication use and increased physiological birth, these outcomes improve with more sessions, although it remains unclear if this trend eventually plateaus [60]. Delivery varied across studies: midwives and nurses led the majority, despite pain education representing less than 1% of programme hours in their training [61]. Conversely, physiotherapy courses included the most pain-related content, suggesting pelvic health physiotherapists may be well-placed to deliver pain-specific education. One study used doulas—non-medical professionals who have demonstrated effectiveness in reducing labour pain, anxiety, and caesarean rates [62]. Despite lacking medical training, doulas may offer a more cost-effective solution and help reduce health disparities [63]. Which profession is best placed to deliver pain-related antenatal education remains unclear and warrants expert consensus.

Analysing pain outcomes highlighted a stark lack of focus on the postpartum period. This may be reflective of the lack of support for women beyond childbirth [64], [6]. In many countries, six weeks after delivery, responsibility for maternal care shifts from the midwife to the woman’s general practitioner or other services. Unfortunately, poor maternal health after this period is either being missed or is costing healthcare systems more because of higher healthcare utilisation for conditions such as persistent postpartum pain and depression, despite many cost-analyses proving the financial worth of screening for such conditions [65], [66]. This demonstrates a current gap in the literature and an opportunity for future researchers to explore the role of antenatal education on persistent postpartum pain outcomes.

Results from this scoping review give an overview of the current methodological quality issues for studies examining the effect of antenatal education on labour and postpartum pain outcomes. Based on the suggested categorisation for the Downs and Black score, all studies scored below the 20-point threshold to indicate high-quality evidence. Most studies were on the lower end of fair methodological quality, while one study was scored as low (13). Some studies did not satisfy certain criteria on the Downs and Black checklist because information was not reported within the publication. The rating criteria that were most commonly unsatisfied in the included studies were related to blinding, randomisation and reporting adverse events.

Many of the included studies employed quasi-experimental or non-randomised controlled designs, reflecting both ethical and logistical barriers to conducting large-scale RCTs in this population. Even among randomised studies, methodological weaknesses were observed. The most consistently unmet checklist items related to blinding of participants and assessors, allocation concealment, and adjustment for confounding variables. These omissions raise concerns about performance, detection, and selection bias.

Furthermore, external validity was universally limited and indicated that intervention settings and delivery staff were not representative of routine antenatal care in each given country. This weakens the generalisability of findings to real-world maternity settings. In addition, power calculations were inadequate in several studies, particularly those with small sample sizes. This may undermine confidence in the reported effect estimates.

Several studies also failed to provide a comprehensive description on whether adverse events were monitored, and compliance with the intervention was often poorly tracked, despite being central to behavioural interventions and clinical trials involving the pregnant women. Only a minority of studies described confounding variables or accounting for losses to follow-up, both of which are critical in evaluating internal validity.

Overall, these methodological limitations suggest that, while many studies reported positive effects of antenatal education interventions, the strength of the evidence is weakened by design flaws, reporting gaps, and inconsistent bias control. Future research should prioritise rigorous randomisation, comprehensive reporting, appropriate statistical adjustments, and assessor blinding to improve the quality and credibility of findings in this field.

### Strengths and limitations

Firstly, this scoping review involved a comprehensive and systematic search across twelve major electronic databases, ensuring a broad capture of relevant literature. To our knowledge, this is the first scoping review to specifically map the content and characteristics of antenatal education programmes focused on pain management through pregnancy, labour, and the postpartum period. The use of rigorous methodology, including adherence to PRISMA-ScR and JBI guidelines, independent and blinded screening by multiple reviewers, and efforts to contact authors for missing data, further enhances the robustness and credibility of the findings. Additionally, the focus on programme content, delivery, dosage, and pain outcomes addresses a critical gap in maternal health education and may help inform future practice and research

One limitation of this study is the reliance on descriptions of antenatal education interventions. Incomplete or inaccurate descriptions by the authors may have distorted or misrepresented the true nature of the interventions. Further, the heterogeneity of study characteristics, meaning inferences about the results may have varied between authors.

Another limitation of the study was that only quantitative studies were included in the review. Including qualitative papers in the review may have captured women’s experiences of pain education and/ or labour. However, only quantitative papers were included to best address the research question.

### Implications for future research

This review highlights a need for high-quality, theory-informed studies that evaluate antenatal education interventions specifically targeting pain management during labour and the postpartum period. Future research should explore the integration of pain neuroscience education tailored to the antenatal population, clearly articulate mechanisms of action, and employ multidimensional outcome measures that capture both sensory and emotional aspects of pain. There is also a pressing need to assess postpartum pain outcomes more comprehensively, investigate the optimal frequency, duration, and mode of delivery of antenatal education, and determine which professionals are best placed to deliver pain-specific content. Establishing consensus on core educational domains through methods such as Delphi studies may further support the development of standardised, evidence-based antenatal education programmes.

## Conclusion

This scoping review aimed to map the contents and structure of antenatal education interventions that target labour and postpartum pain outcomes. Findings showed antenatal education topics included limited content on pain management and lacked consistency across interventions. The most common topic was the distinction between “true and false labour pain”, with true indicating regular contractions progressing to labour, and false indicating irregular contractions not leading to labour. Only one study included the topic of pain physiology. Postpartum pain outcomes were also under-addressed. The findings of this review add to the growing need for targeted interventions that address labour and postpartum pain management through non-pharmacological methods. An understanding of pain mechanisms, including the neurobiology and biopsychosocial aspects of pain, has been shown in other populations mitigate pain-catastrophising, reduce fear-avoidance, and improve self-efficacy in managing painful experiences. Therefore, we suggest that future antenatal education programmes explore incorporating PNE principles. By providing pregnant individuals with an evidence-based understanding of pain, PNE could promote self-efficacy and reduce pain-related distress. Further research is warranted to develop, implement, and evaluate PNE interventions specifically tailored for antenatal education settings.

## Data Availability

All relevant data are within the manuscript and its Supporting Information files.

## Acknowledgements

This work was supported by the National Institute for Health Research INSIGHT Programme, in conjunction with Bournemouth University. The views expressed in this publication are those of the authors and not necessarily those directly of the National Institute for Health and Care Research or the Department of Health and Social Care.

